# An external validation of the QCovid risk prediction algorithm for risk of mortality from COVID-19 in adults: national validation cohort study in England

**DOI:** 10.1101/2021.01.22.21249968

**Authors:** Vahe Nafilyan, Ben Humberstone, Nisha Mehta, Ian Diamond, Carol Coupland, Luke Lorenzi, Piotr Pawelek, Ryan Schofield, Jasper Morgan, Paul Brown, Ronan Lyons, Aziz Sheikh, Julia Hippisley-Cox

## Abstract

**Background:** To externally validate a risk prediction algorithm (QCovid) to estimate mortality outcomes from COVID-19 in adults in England.

**Methods:** Population-based cohort study using the ONS Public Health Linked Data Asset, a cohort based on the 2011 Census linked to Hospital Episode Statistics, the General Practice Extraction Service Data for pandemic planning and research, radiotherapy and systemic chemotherapy records. The primary outcome was time to COVID-19 death, defined as confirmed or suspected COVID-19 death as per death certification. Two time periods were used: (a) 24^th^ January to 30^th^ April 2020; and (b) 1^st^ May to 28^th^ July 2020. We evaluated the performance of the QCovid algorithms using measures of discrimination and calibration for each validation time period.

**Findings:** The study comprises 34,897,648 adults aged 19-100 years resident in England. There were 26,985 COVID-19 deaths during the first time-period and 13,177 during the second. The algorithms had good calibration in the validation cohort in both time periods with close correspondence of observed and predicted risks. They explained 77.1% (95% CI: 76.9% to 77.4%) of the variation in time to death in men in the first time-period (R^2^); the D statistic was 3.76 (95% CI: 3.73 to 3.79); Harrell’s C was 0.935 (0.933 to 0.937). Similar results were obtained for women, and in the second time-period. In the top 5% of patients with the highest predicted risks of death, the sensitivity for identifying deaths in the first time period was 65.9% for men and 71.7% for women. People in the top 20% of predicted risks of death accounted for 90.8% of all COVID-19 deaths for men and 93.0% for women.

**Interpretation:** The QCovid population-based risk algorithm performed well, showing very high levels of discrimination for COVID-19 deaths in men and women for both time periods. It has the potential to be dynamically updated as the pandemic evolves and therefore, has potential use in guiding national policy.

**Funding:** National Institute of Health Research

**RESEARCH IN CONTEXT:** *Evidence before this study:* Public policy measures and clinical risk assessment relevant to COVID-19 need to be aided by rigorously developed and validated risk prediction models. A recent living systematic review of published risk prediction models for COVID-19 found most models are subject to a high risk of bias with optimistic reported performance, raising concern that these models may be unreliable when applied in practice. A population-based risk prediction model, QCovid risk prediction algorithm, has recently been developed to identify adults at high risk of serious COVID-19 outcomes, which overcome many of the limitations of previous tools.

*Added value of this study:* Commissioned by the Chief Medical Officer for England, we validated the novel clinical risk prediction model (QCovid) to identify risks of short-term severe outcomes due to COVID-19. We used national linked datasets from general practice, death registry and hospital episode data for a population-representative sample of over 34 million adults. The risk models have excellent discrimination in men and women (Harrell’s C statistic>0.9) and are well calibrated. QCovid represents a new, evidence-based opportunity for population risk-stratification.

*Implications of all the available evidence:* QCovid has the potential to support public health policy, from enabling shared decision making between clinicians and patients in relation to health and work risks, to targeted recruitment for clinical trials, and prioritisation of vaccination, for example.

## INTRODUCTION

The first cases of SARS-CoV-2 infection were reported in the United Kingdom (UK) on the 24^th^ January 2020, with the first COVID-19 death on the 28^th^ February 2020. As of 19 January 2021, there have been over 90,000 deaths from COVID-19 in the UK, and over 2 million deaths globally^1^.

Emerging evidence throughout the course of the pandemic, initially from case series, and then cohorts of individuals with confirmed SARS-CoV-2 infection, has demonstrated associations of age, sex, certain co-morbidities, ethnicity and obesity with adverse COVID-19 outcomes such as hospitalisation and death^2-9^. There is now a growing knowledge base regarding risk factors for severe COVID-19. As many countries are re-introducing ‘lockdown’ measures and vaccination programmes have started being rolled out, there is an opportunity to develop more nuanced guidance^10^ based on predictive algorithms to inform risk management decisions. Better knowledge of individuals’ risks could also help guide decisions on managing occupational risk and in targeting of vaccines to those most at risk. Whilst several risk prediction models have been developed, a recent systematic review found that they all have high risk of bias and that their reported performance is optimistic^11^.

The use of primary care datasets such as QResearch with linkage to registries such as death records and hospital admissions data represents an innovative approach to clinical risk prediction modelling for COVID-19 which has successfully been developed, validated, and implemented in the NHS over the last 10 years^12-14^. It provides accurately coded, individual-level data for very large numbers of people representative of the national population. This approach was used to develop the QCovid prediction models^15^ drawing on the rich phenotyping of individuals with demographic, medical and pharmacological predictors to allow robust statistical modelling and evaluation. Such linked datasets have a track record for the development, and evaluation of established clinical risk models including for cardiovascular disease^12^, diabetes^14^ and mortality^13^. Whilst QCovid predicts both COVID- 19 hospital related admission and COVID-19 death, the aim of this analysis is to validate the mortality outcome which estimate the risks of becoming infected and subsequent death due to COVID-19 in an extremely large national cohort. A companion study currently underway will externally validate these, using datasets from Wales using ^16^SAIL^17^ and Scotland using EAVE-II^18^ and will be reported separately.

## METHODS

The Chief Medical Officer for England asked the New and Emerging Respiratory Virus Threats Advisory Group (NERVTAG), to develop and validate a clinical risk prediction model for COVID-19 in line with the emerging evidence. The resulting QCovid model was developed and validated using the QResearch database and reported in accordance with TRIPOD^19^ and RECORD^20^ guidelines and with input from a patient advisory group. The original protocol is published ^21^ along with the results of the paper reporting the original derivation and validation of the model^15^. This paper reports the validation of the model on an independent data source.

### Study design and data sources

We undertook a validation cohort study of individuals aged 19-100 years using the Office for National Statistics (ONS) Public Health Linked Data Asset. This dataset is based on the 2011 Census in England, linked at individual level using the NHS number to mortality records, Hospital Episode Statistics and the General Practice Extraction Service (GPES) Data for pandemic planning and research. To obtain NHS numbers, the 2011 Census was linked to the 2011-2013 NHS Patient Registers using deterministic and probabilistic matching, with an overall linkage rate of 94.6%. We excluded patients (approximately 13.1%) who did not have a valid NHS number. For the purpose of the validation of the QCovid algorithm, we further linked radiotherapy and systemic chemotherapy records based on NHS number. The ONS Public Health Linked Data Asset includes data on most patients used to develop the QCovid algorithm, but also includes patients registered with practices using IT systems other than EMIS, such as TPP (used by 35% of GP practices).

We identified a cohort of all individuals aged 19-100 years who were enumerated at the 2011 Census and registered alive and resident in England on 24^th^ January 2020. Patients entered the cohort on 24^th^ January 2020 (date of first confirmed COVID-19 case in UK) and were followed up until they had the outcome of interest or 28^th^ July 2020, which is the date up to which linked data were available at the time of the analysis. This also extends the period of observation beyond the original QCovid study. We divided the study period into two time periods: 24^th^ January 2020 to 30^th^ April 2020 and 01 May 2020 to 28^th^ July 2020.

### Outcomes

The outcome was time to COVID-19 death (either in hospital or out of hospital), defined as confirmed or suspected COVID-19 death as identified by two ICD10 codes (U07.1 or U07.2) recorded on the death certification.

### Predictor variables

We derived pre-existing conditions and demographic characteristics using the same definitions as used to develop the QCovid algorithm. The primary care records used in the ONS Public Health Linked Data Asset were based on an existing GPES dataset which included many but not all of the relevant clinical codes used to develop the QCovid algorithm. Nonetheless, we derived most of the pre-existing conditions, although we could not identify patients who had a solid organ or bone marrow transplant in the past six months; those on kidney dialysis; those with sickle cell disease or severe combined immune deficiency syndrome; Similarly, we could not distinguish between patients suffering from type 1 or type 2 diabetes. Variables used to validate the QCovid algorithm are listed in Box 1.

### Model validation

We fitted an imputation model to replace missing values for body mass index (BMI), using predicted values from linear regression models stratified by sex. Predictors included all predictor variables in the QCovid algorithm, interacted with age.

We applied the QCovid risk equations (version 1) to men and women in the validation dataset and evaluated R^2^ values^**22**^, Brier scores and measures of discrimination and calibration^**23** 24^ with corresponding 95% confidence intervals (CIs) over the two time periods. R^2^ values refer to the proportion of variation in survival time explained by the model. Brier scores measure predictive accuracy where lower values indicate better accuracy^25^. The D statistic is a discrimination measure which quantifies the separation in survival between patients with different levels of predicted risks and the Harrell’s C-statistics is a discrimination metric which quantifies the extent to which people with higher risk scores have earlier events. Model calibration was assessed in the two time periods by comparing mean predicted risks with observed risks by twentieths of predicted risk. Observed risks were derived in each of the 20 groups using non-parametric estimates of the cumulative incidences.

The performance metrics were calculated in the whole cohort and in the following pre-specified subgroups: 5-year age-sex bands, nine ethnic groups, and within each quintile of the Townsend Index, a measure of deprivation. We also estimated the performance metrics on a sample restricted to patients registered with practices using the TPP system, and therefore not used at all to derive the algorithm.

### Ethics

The ethics approval for the development and validation of QCovid was granted by the East Midlands-Derby Research Ethics Committee [reference 18/EM/0400].

### Role of the funding source

This study is funded by a grant from the National Institute for Health Research following a commission by the Chief Medical Officer for England whose office contributed to the development of the study question and facilitated access to relevant national datasets, contributed to interpretation of data, drafting of the report.

## RESULTS

### Overall study population

Overall, 34,897,648 people in England aged 19-100 years met our inclusion criteria. Out of the 40,136,597 people aged 19-100 who were enumerated at the 2011 Census and were alive on 24^th^ January 2020, 5,238,949 (13.1%) people were excluded because they did not have a valid NHS number or had an NHS number that could not be linked to the GPES data. This could be because they migrated out of England, and therefore were no longer registered with the NHS in England. Our data cover 80.0% of the population in England aged 19 or over (See Supplementary Table 1). The coverage is lowest in London (68.2%) and highest in Yorkshire & Humber (83.7%).

### Baseline characteristics

Table 1 shows the baseline characteristics of patients. Of these patients, 16,599,875 (47.57%) were men and 6,052,563 (17.34%) were of ethnic minority background. The mean age was 51.1 years, which is slightly higher than in the cohort used to derive the QCovid models (48.2 years). For most pre-existing conditions, the estimated prevalence in the ONS Public Health Linked Data Asset is similar to the prevalence in the QResearch derivation cohort. However, the ONS dataset had higher proportions of people taking anti-leukotriene or long acting beta2-agonists, or being prescribed oral steroids in the last six months because of data limitations.

**Table 1:** Demographic and medical characteristics for the validation cohort and those who died with COVID-19 in the two time periods. Results are numbers (column %) except where otherwise specified.

|  | **cohort total** | **Period 1** | **Period 2** |
| --- | --- | --- | --- |
|  |  | **24.01.2020-30.04.2020** | **01.05.2020-31.07.2020** |
| **Total** | 34,897,648 | 26,985 | 13,177 |
| Females | 18,297,773 (52.43) | 11,651 (43.18) | 6560 (49.78) |
| Males | 16,599,875 (47.57) | 15,334 (56.82) | 6,617 (50.22) |
| mean age (SD) | 51.09 (18.76) | 79.98 (11.63) | 82.13 (10.79) |
| **Age-band** |  |  |  |
| 19-29 years | 5,601,475 (16.05) | 44 (0.16) | 13 (0.10) |
| 30-39 years | 5,268,030 (15.10) | 116 (0.43) | 30 (0.23) |
| 40-49 years | 5,625,225 (16.12) | 364 (1.35) | 125 (0.95) |
| 50-59 years | 6,435,204 (18.44) | 1,196 (4.43) | 400 (3.04) |
| 60-69 years | 5,185,917 (14.86) | 2,727 (10.11) | 962 (7.30) |
| 70-79 years | 4,225,729 (12.11) | 6,280 (23.27) | 2,695 (20.45) |
| 80-89 years | 2,093,545 (6.00) | 10,841 (40.17) | 5,580 (42.35) |
| 90+ years | 462,523 (1.33) | 5,417 (20.07) | 3,372 (25.59) |
| **Geographical region** |  |  |  |
| East Midlands | 3,137,521 (8.99) | 1,979 (7.33) | 1,372 (10.41) |
| East of England | 3,987,067 (11.43) | 2,549 (9.45) | 1,456 (11.05) |
| London | 4,662,731 (13.36) | 5,403 (20.02) | 956 (7.26) |
| North East | 1,755,316 (5.03) | 1,429 (5.30) | 931 (7.07) |
| North West | 4,643,947 (13.31) | 4,289 (15.89) | 2,411 (18.30) |
| South East | 5,818,470 (16.67) | 4,005 (14.84) | 2,118 (16.07) |
| South West | 3,674,549 (10.53) | 1,657 (6.14) | 745 (5.65) |
| West Midlands | 3,643,447 (10.44) | 3,284 (12.17) | 1,497 (11.36) |
| Yorkshire & Humber | 3,574,600 (10.24) | 2,390 (8.86) | 1,691 (12.83) |
| **Ethnicity** |  |  |  |
| Bangladeshi | 258,053 (0.74) | 179 (0.66) | 29 (0.22) |
| Black African | 520,547 (1.49) | 398 (1.47) | 62 (0.47) |
| Black Caribbean | 374,982 (1.07) | 732 (2.71) | 124 (0.94) |
| Chinese | 185,966 (0.53) | 107 (0.40) | 27 (0.20) |
| Indian | 931,247 (2.67) | 800 (2.96) | 216 (1.64) |
| Mixed | 551,567 (1.58) | 184 (0.68) | 67 (0.51) |
| Other | 835,506 (2.39) | 590 (2.19) | 130 (0.99) |
| Pakistani | 679,062 (1.95) | 426 (1.58) | 123 (0.93) |
| White British | 28,845,085 (82.66) | 22,462 (83.24) | 12,018 (91.20) |
| White other | 1,715,633 (4.92) | 1,107 (4.10) | 381 (2.89) |
| **Townsend deprivation quintile** |  |  |  |
| 1 (most affluent) | 7,491,652 (21.47) | 4,993 (18.50) | 2,842 (21.57) |
| 2 | 7,738,292 (22.17) | 5,326 (19.74) | 2,967 (22.52) |
| 3 | 6,834,804 (19.58) | 5,111 (18.94) | 2,647 (20.09) |
| 4 | 6,467,204 (18.53) | 5,365 (19.88) | 2,472 (18.76) |
| 5 (most deprived) | 6,366,096 (18.24) | 6,190 (22.94) | 2,249 (17.07) |
| **Accommodation** |  |  |  |
| Neither homeless or care home | 34,667,007 (99.34) | 19,995 (74.10) | 9,039 (68.60) |
| Care home or nursing home | 230,641 (0.66) | 6,990 (25.90) | 4,138 (31.40) |
| **Body mass index (kg/m<sup>2</sup>)** |  |  |  |
| BMI < 18.5 | 393,928 (1.13) | 983 (3.64) | 614 (4.66) |
| BMI 18.5-24.99 | 6,658,276 (19.08) | 5,776 (21.40) | 2,965 (22.50) |
| BMI 25-29.99 | 6,661,721 (19.09) | 5,552 (20.57) | 2,385 (18.10) |
| BMI 30+ | 5,661,007 (16.22) | 5,540 (20.53) | 2,066 (15.68) |
| BMI not recorded | 15,522,716 (44.48) | 9,134 (33.85) | 5,147 (39.06) |
| **Chronic kidney disease (CKD)** |  |  |  |
| no CKD | 34,392,544 (98.55) | 24,425 (90.51) | 11,939 (90.60) |
| CKD3 | 436,595 (1.25) | 1,820 (6.74) | 914 (6.94) |
| CKD4 | 45,638 (0.13) | 452 (1.68) | 205 (1.56) |
| CKD5 | 22,871 (0.07) | 288 (1.07) | 119 (0.90) |
| **Learning disability:** |  |  |  |
| No learning disability | 34,393,288 (98.55) | 25,300 (93.76) | 12,386 (94.00) |
| Learning disability | 490,357 (1.41) | 1,616 (5.99) | * |
| Down's Syndrome | 14,003 (0.04) | 69 (0.26) | * |
| **Chemotherapy:** |  |  |  |
| No chemotherapy in last 12 months | 34,776,317 (99.65) | 26,472 (98.10) | 12,908 (97.96) |
| Chemotherapy group A | 38,956 (0.11) | 128 (0.47) | 62 (0.47) |
| Chemotherapy group B | 76,763 (0.22) | 339 (1.26) | 180 (1.37) |
| Chemotherapy group C | 5,612 (0.02) | 46 (0.17) | 27 (0.20) |
| **Cancer and immunosuppression:** |  |  |  |
| Blood cancer | 336,990 (0.97) | 897 (3.32) | 465 (3.53) |
| Respiratory cancer | 9,720 (0.03) | 142 (0.53) | 66 (0.50) |
| Radiotherapy in last 6 months | 56,252 (0.16) | 174 (0.64) | 100 (0.76) |
| Solid organ transplant | 3,488 (0.01) | 26 (0.10) | * |
| Prescribed immunosuppressant medication by GP | 7,237 (0.02) | 20 (0.07) | * |
| Prescribed leukotriene or LABA | 2,362,855 (6.77) | 4,956 (18.37) | 2,319 (17.60) |
| prescribed regular prednisolone | 404,467 (1.16) | 2,124 (7.87) | 1,028 (7.80) |
| **Other co-morbidities** |  |  |  |
| Diabetes | 3,087,792 (8.85) | 8,700 (32.24) | 3,650 (27.70) |
| COPD | 1,053,783 (3.02) | 3,814 (14.13) | 1,809 (13.73) |
| asthma | 4,382,954 (12.56) | 3,344 (12.39) | 1,504 (11.41) |
| Rare pulmonary diseases | 373,807 (1.07) | 1,707 (6.33) | 734 (5.57) |
| Pulmonary hypertension or pulmonary fibrosis | 127,760 (0.37) | 1,158 (4.29) | 502 (3.81) |
| Coronary heart disease | 1,549,243 (4.44) | 5,946 (22.03) | 2,861 (21.71) |
| Stroke | 902,277 (2.59) | 5,086 (18.85) | 2,685 (20.38) |
| Atrial Fibrillation | 1,096,209 (3.14) | 5,237 (19.41) | 2,894 (21.96) |
| Congestive cardiac failure | 545,617 (1.56) | 3,739 (13.86) | 1,830 (13.89) |
| Venous thromboembolism | 8,878 (0.03) | 35 (0.13) | * |
| Peripheral vascular disease | 303,118 (0.87) | 1,588 (5.88) | 771 (5.85) |
| Congenital heart disease | 359 (0.00) | * | 0 (0.00) |
| Dementia | 414,540 (1.19) | 8,293 (30.73) | 4,699 (35.66) |
| Parkinson's disease | 113,647 (0.33) | 1,021 (3.78) | 573 (4.35) |
| Epilepsy | 405,047 (1.16) | 797 (2.95) | 387 (2.94) |
| Rare neurological conditions | 27,583 (0.08) | 149 (0.55) | 48 (0.36) |
| Cerebral palsy | 4,350 (0.01) | 31 (0.11) | * |
| Severe mental illness | 6,574,526 (18.84) | 5,341 (19.79) | 2,541 (19.28) |
| Osteoporotic fracture | 29,153 (0.08) | 194 (0.72) | 92 (0.70) |
| Rheumatoid arthritis or SLE | 315,431 (0.90) | 696 (2.58) | 369 (2.80) |
| Cirrhosis of the liver | 81,753 (0.23) | 241 (0.89) | 114 (0.87) |

\* represents values which have been suppressed due to small numbers < 5

26,985 (0.08%) patients had a COVID-19 related death during the first period: 24 January 2020 to 30 April 2020). 13,177 (0.04%) patients had a COVID-19 related death during the second period (1 May 2020 to 28 July 2020). Out of the 49,461 deaths that occurred in England over the period, 81.2% of these are included in our data (See Supplementary Table 1). The coverage is lowest in London (74.2%) and highest in the North West (84.6%). In both periods, COVID-19 deaths occurred across all regions, with the greatest numbers in London in period 1, (5,403 - 20.02% of all deaths) and in the North West in period 2 (2,411 - 18.30%). Of those who died in period 1, 15,334 (56.82%) were male; 4523 (16.76%) were from ethnic minority groups; 22,538 (83.52%) were aged 70 and over; 8,700 (32.24%) had diabetes; 8,293 (30.73%) had dementia; 6,990 (25.90%) were identified as living in a care home. Those who had a COVID-19 related death in period 2 had a similar profile to those in period 1 but were on average older (88.4% aged 70 and over) and more likely to live in a care home (31.4%).

### Discrimination

Table 2 shows the performance of the risk equations in the validation cohort for women and men in the two time periods. Overall, the values for the R^2^, D and C statistics were high and similar in women and men in both periods. In the first period for women, the equation explained 76.3% of the variation in time to COVID-19 death; Harrell’s C statistic was 0.945 and the D statistic 3.67. The corresponding values in men were 77.1%, 0.935 and 3.76 respectively. All these discrimination metrics are higher than in the original QResearch cohort used to validate the algorithm. The results were similar for the second validation period. In period 2 for women, the R^2^ was 75.4%, Harrell’s C statistic 0.956, the D statistic 3.58. The corresponding values for men were 77.4%, 0.944 and 3.78 respectively. Similar results were obtained when restricting the sample to 14,104,452 patients registered with practices using the TPP system (Supplementary Table 2).

**Table 2.** Performance of the risk models to predict risk of COVID-19 death in the validation cohort.

|  | **Period 1 (24/01/2020 -30/04/2020)** |  | **Period 2 (01/05/2020 -28/07/2020)** |  |
| --- | --- | --- | --- | --- |
|  | **COVID -19 death** | **COVID -19 death** | **COVID -19 death** | **COVID -19 death** |
|  | **females** | **males** | **females** | **males** |
|  | Estimate (95% CI) | Estimate (95% CI) | Estimate (95% CI) | Estimate (95% CI) |
| R<sup>2</sup> statistic | 0.763<br>(0.760 to 0.766) | 0.771<br>(0.769 to 0.774) | 0.754<br>(0.750 to 0.757) | 0.774<br>(0.769 to 0.777) |
| D statistic | 3.671<br>(3.640 to 3.702) | 3.761<br>(3.732 to 3.789) | 3.579<br>(3.542 to 3.616) | 3.782<br>(3.739 to 3.826) |
| Harrell's C statistic | 0.945<br>(0.943 to 0.947) | 0.935<br>(0.933 to 0.937) | 0.956<br>(0.954 to 0.958) | 0.944<br>(0.942 to 0.946) |
| Brier score | 0.0018 | 0.0013 | 0.0007 | 0.0008 |

Figure 1 displays Harrell’s C statistic by age-band for men and women in period 1 (Panel A) and period 2 (Panel B). The Harrell’s C statistics are over 0.7 for all age bands indicating that even within each age band the model discriminates well. The C statistics are lower for patients aged 90 or over than for younger patients. The C statistics, R^2^, and D by age-band, deprivation quintile and ethnic group in men and women for both periods are reported in Supplementary Tables 3, 4 and 5. Performance was generally similar to the overall results except for age where the performance was lower within individual age-bands.

**Figure 1.**
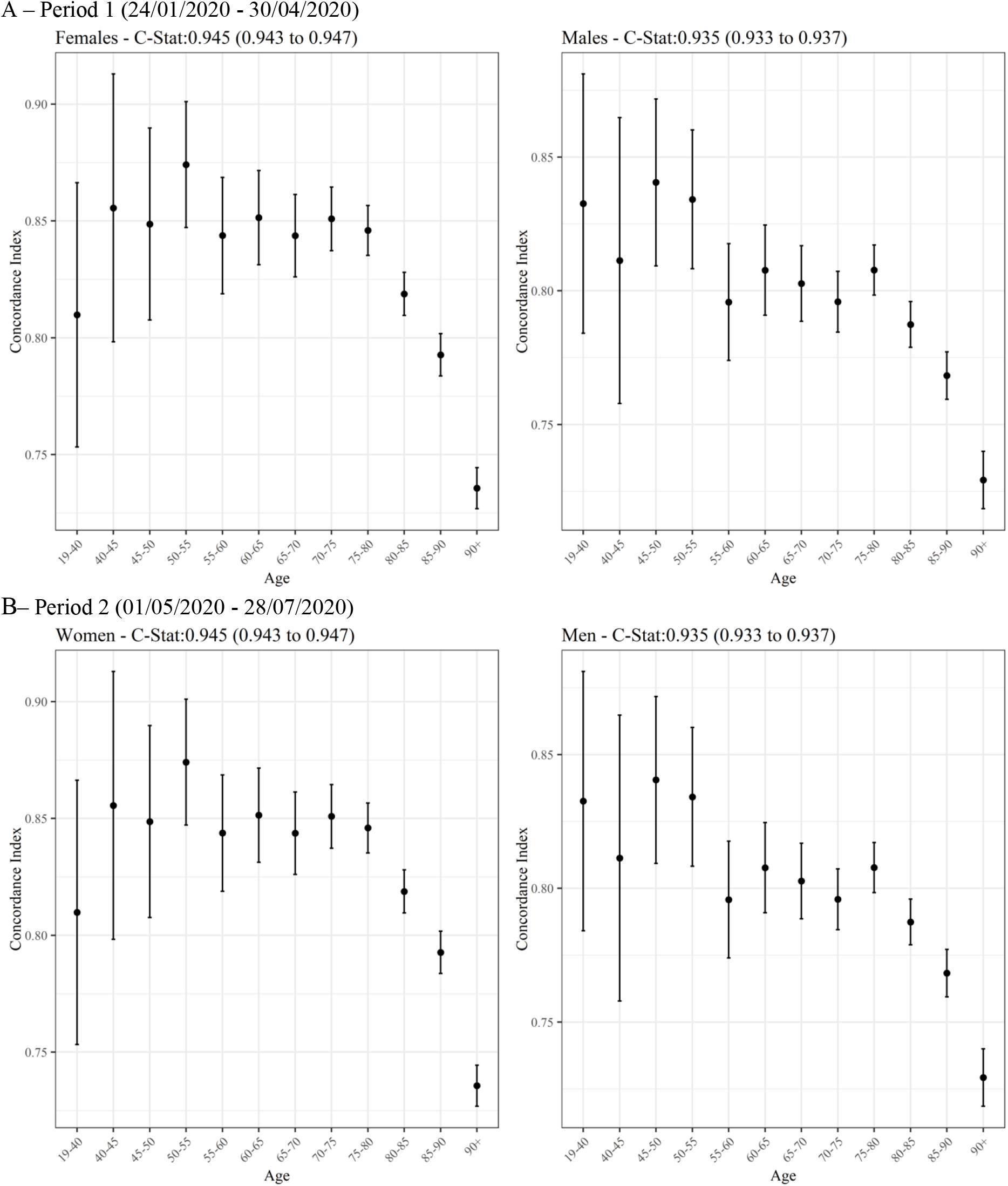

### Calibration

Figure 2 displays the calibration plots for the COVID-19 mortality equation for men and women and in both periods. Overall, both sets of equations were well calibrated, as the predicted and observed risks were similar. However, as in the original QResearch validation cohort, the model underestimates the risk of COVID-19 death for those in the top 5% of the predicted risk score.

**Figure 2:**
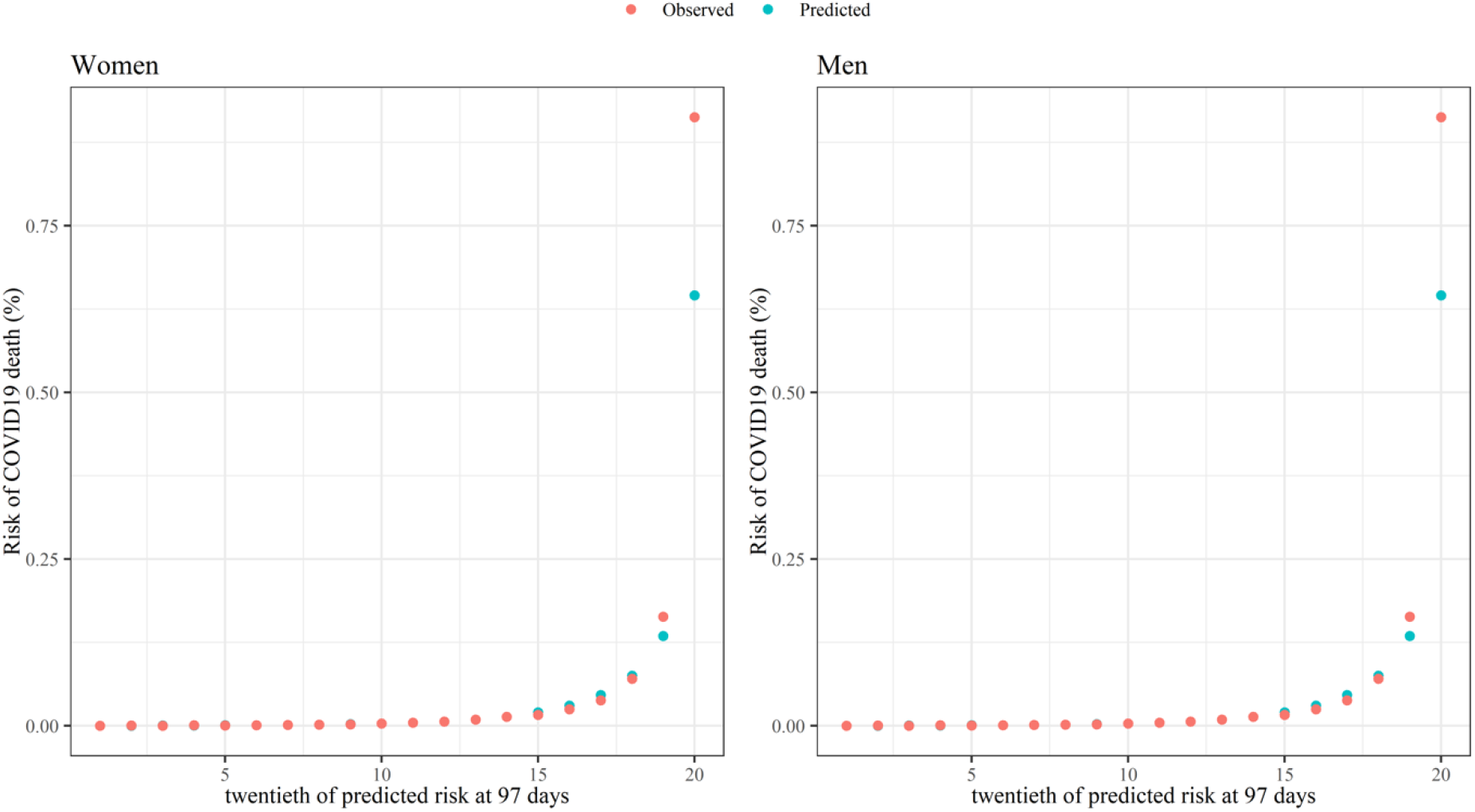
Predicted and observed risk of covid-19 related death in first study period (24 January to 30 April 2020)

### Risk stratification

Figure 3 shows the sensitivity values for the mortality equation in period 1 (24 January 2020 – 30 April 2020) and period 2 (1 May 2020 – 28 July 2020) evaluated at different thresholds based on the centiles of the predicted absolute risk in the validation cohort. Full results are reported in Supplementary Table 6. The sensitivity was higher in women than in men, and in period 2 than period 1. In period 1, 65.94% of deaths in men occurred in those in the top 5% for predicted absolute risk of death from Covid-19 (90 day predicted absolute risks above 0.289%) and 71.67% of deaths in women occurred in the top 5% (predicted absolute risks above 0.188%). In period 2, 71.10% of deaths occurred in men in the top 5% for predicted absolute risk of death from Covid-19 (predicted absolute risks above 0.278%) and 77.16% of deaths occurred in women in the top 5% (predicted absolute risks above 0.181%). Supplementary Figure 1 shows the sensitivity for the two time periods based on relative risks (defined as the ratio of the individual’s predicted absolute risk to the predicted absolute risk for a person of the same age and sex with a white ethnicity, body mass index of 25, and mean deprivation score with no other risk factors). In period 1, 40.56% of deaths occurred in men in the top 5% for predicted relative risk of death from Covid-19, and 42.63% for women. In period 2, 42.62% of deaths occurred in men in the top 5% for predicted relative risk of death from Covid-19, and 43.57% for women.

**Figure 3:**
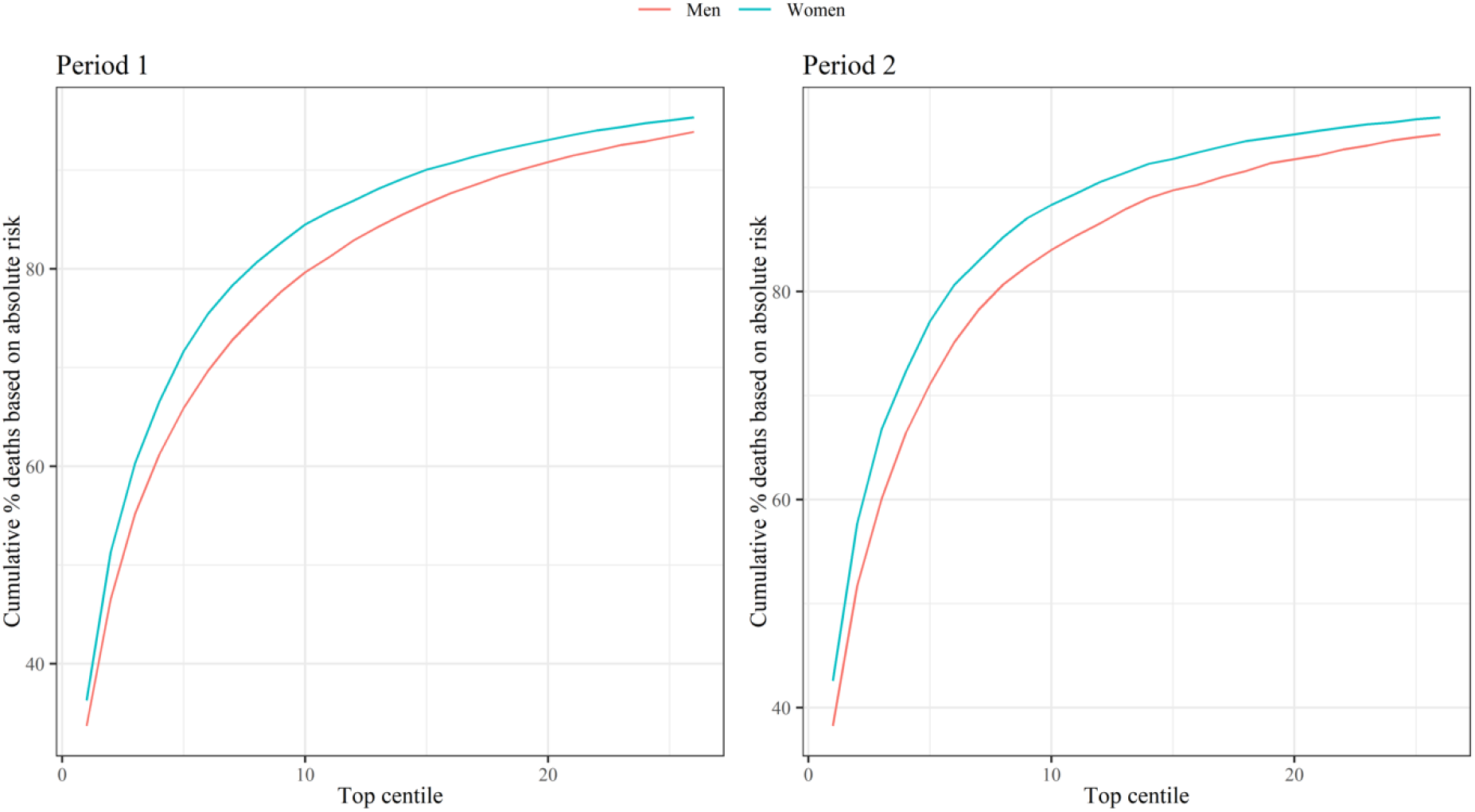
Sensitivity for covid-19 related death in validation cohort for period 1 (24 January 2020 – 30 April 2020) and period 2 (1 May 2020 – 28 July 2020) Centiles based on predicted absolute risks in men and women in each period. Sensitivity (cumulative % deaths) is percentage of total deaths in the period that occurred within the group of patients above the predicted risk threshold.

## DISCUSSION

We have validated the QCovid clinical risk prediction model for mortality due to COVID-19 using a national external linked dataset. We have used national linked datasets from the 2011 Census, general practice, death registry data for a population-representative sample of nearly 35 million adults. The risk models have excellent discrimination (Harrell’s C statistics>0.9), are well calibrated and have a high sensitivity.

Our study had a number of important strengths. First, we used a unique linked dataset based on the 2011 Census for nearly 35 million people living in England. Second, we used a wide range of metrics, over two time periods to validate the QCovid predictive model, extending the period of observation beyond the original study. All the metrics in the two time periods for both men and women indicate that the algorithm performs well, and the metrics are comparable with the original validation of QCovid in the QResearch database^15^. Finally, we showed that the model’s performance was similar when restricting the sample to patients that were registered with practices using a different clinical computer system provider (TPP), and therefore not used to derive the QCovid model.

This study also has several limitations. First, because of data limitations, we could not derive all predictors in the same way as in the derivation cohort. Despite these inconsistencies, the model had excellent discrimination and calibration. Second, we only focused on COVID-19 related deaths, but not hospital admissions, because of the lack of data. Finally, because the Public Health Data Asset is based on the 2011 Census, our sample was restricted to patients who were enumerated in 2011, that is about 94% of the population living in England in 2011. Recent migrants were excluded from this study, but they tend to be younger than the native population and therefore at lower risk of COVID-19 death.

QCovid represents a new approach for population risk-stratification for adverse outcomes from COVID-19, and our validation indicates that the risk algorithm performs well on external data not used for its derivation. Whilst it has been specifically designed to inform UK health policy and interventions to manage COVID-19 related risks, it also has international potential, subject to local validation. It could also be deployed in a number of health and care applications, either during the current phase of the pandemic, or in subsequent ‘waves’ of infection. These could include supporting targeted recruitment for clinical trials, vaccine prioritisation, and discussions between patients and clinicians in relation to work and health risks, for example through weight reduction since obesity is the single most important modifiable risk factor for serious COVID-19 complications^8^.

In conclusion, this study presents a robust validation of a new prediction model that could be used to support population risk stratification in relation to public health interventions, for example vaccine utilisation. We anticipate that the algorithms will be updated regularly as understanding of COVID-19 increases, as more data become available, as new variants emerge, effective treatments for COVID become available, the vaccination program rolls-out, immunity levels change or as behaviour in the population changes and hence we anticipate that this validation will need to be repeated on a regular basis. It is important for patients/carers, and clinicians that there is a common appropriately developed evidence-based model that is consistently implemented and is supported by the academic, clinical and patient communities. This will then help ensure consistent policy and clear national communication between policy makers, professionals, employers and the public.

## Data Availability

The ONS Public Health Linked Data Asset will be made available on the ONS Secure Research Service for Accredited researchers. Researchers can apply for accreditation through the Research Accreditation Service. The data will include all variables used in this analysis, except predictors based on radiotherapy and systemic chemotherapy records, which cannot be shared.

## Acknowledgements

We acknowledge the contribution Jenny Harries, Nazmus Haq, Joanna Moody and Shamim Rahman from the UK Department of Health and Social Care, Joy Preece and Dan Ayoubkhani from the Office for National Statistics. This project involves data derived from patient-level information collected by the NHS, as part of the care and support of cancer patients. Access to the data was facilitated by the PHE Office for Data Release. The Hospital Episode Statistics data used in this analysis are re-used by permission from NHS Digital who retain the copyright for these data.

## Ethics approval

Ethics approval for the development and validation of QCovid is covered by Research Ethics Committee [reference 03/4/021].

## Authors and contributors

Study conceptualisation was led by NM, JHC, VN, CC. All authors contributed to the development of the research question, study design, with development of advanced statistical aspects led by CC, VN. VN, RS, PB, PP, JHC and JM, were involved in data specification, curation and collection. JHC developed, checked or updated clinical code groups. VN led the statistical analyses which were checked by LL. All authors contributed to the interpretation of the results. VN and JHC wrote the first draft of the paper. All authors contributed to the critical revision of the manuscript for important intellectual content and approved the final version of the manuscript.

VN had full access to all data in the study and takes responsibility of the integrity of the data and the accuracy of the data analysis. The lead author (VN) affirms that the manuscript is an honest, accurate, and transparent account of the study being reported; that no important aspects of the study have been omitted; and that any discrepancies from the study as planned have been explained.

## Declarations of interest

All authors have completed the ICMJE uniform disclosure form at www.icmje.org/coi_disclosure.pdf and declare: JHC reports grants from National Institute for Health Research Biomedical Research Centre, Oxford, grants from John Fell Oxford University Press Research Fund, grants from Cancer Research UK (CR-UK) grant number C5255/A18085, through the Cancer Research UK Oxford Centre, grants from the Oxford Wellcome Institutional Strategic Support Fund (204826/Z/16/Z), during the conduct of the study. JHC is an unpaid director of QResearch, a not-for-profit organisation which is a partnership between the University of Oxford and EMIS Health who supply the QResearch database used for this work. JHC is a founder and shareholder of ClinRisk ltd and was its medical director until 31^st^ May 2019. ClinRisk Ltd produces open and closed source software to implement clinical risk algorithms (outside this work) into clinical computer systems.

## Box 1

**Predictor variables used to validate the QCOVID model**

- Age in years (continuous)
- Townsend deprivation score (continuous)
- Accommodation (Neither homeless nor care home, Care home or nursing home)
- Ethnicity in nine categories (Bangladeshi, Black African, Black Caribbean, Chinese, Indian, Mixed, Pakistani, White British, White other, Other)
- Body Mass Index (kg/m2)
- Chronic kidney disease (CKD) - (no CKD, CKD3, CKD4, CKD5)
- Learning disability (No learning disability, Down’s Syndrome, other learning disability)
- Chemotherapy in last 12 months (Chemotherapy group A, B, C)
- Respiratory cancer
- Radiotherapy in last 6 months
- Solid organ transplant
- Prescribed immunosuppressant medication by GP
- Prescribed leukotriene or long-acting beta blockers
- Prescribed regular prednisolone
- Sickle cell disease
- Diabetes
- Chronic obstructive pulmonary disease (COPD)
- Asthma
- Rare pulmonary diseases
- Pulmonary hypertension or pulmonary fibrosis
- Coronary heart disease
- Stroke
- Atrial Fibrillation
- Congestive cardiac failure
- Venous thromboembolism
- Peripheral vascular disease
- Congenital heart disease
- Dementia
- Parkinson’s disease
- Epilepsy
- Rare neurological conditions
- Cerebral palsy
- Severe mental illness (bipolar disorder, schizophrenia, severe depression)
- Osteoporotic fracture
- Rheumatoid arthritis or Systemic lupus erythematosus
- Cirrhosis of the liver

Note: For the validation of the QCovid risk model, all patients with diabetes were assigned the coefficient type 2 diabetes. Patients with Stage 5 chronic kidney disease (CKD) were assigned the coefficient for CKD without transplant nor dialysis.

## Appendix

**Supplementary Table 1:** Population and Covid-19 deaths in the ONS Public Health Data Asset compared to England (up to 28^th^ July 2020)

|  | Population |  |  | COVID-19 deaths |  |  |
| --- | --- | --- | --- | --- | --- | --- |
|  | In the Cohort | England | Proportion | Recorded in the Cohort | Recorded in England | Proportion |
| East Midlands | 3,137,521 | 3,779,186 | 83.0% | 3,351 | 3,977 | 84.3% |
| East of England | 3,987,067 | 4,822,148 | 82.7% | 4,005 | 5,113 | 78.3% |
| London | 4,662,731 | 6,834,636 | 68.2% | 6,359 | 8,570 | 74.2% |
| North East | 1,755,316 | 2,108,996 | 83.2% | 2,360 | 2,834 | 83.3% |
| North West | 4,643,947 | 5,696,784 | 81.5% | 6,700 | 7,923 | 84.6% |
| South East | 5,818,470 | 7,107,605 | 81.9% | 6,123 | 7,374 | 83.0% |
| South West | 3,674,549 | 4,457,165 | 82.4% | 2,402 | 2,916 | 82.4% |
| West Midlands | 3,643,447 | 4,566,619 | 79.8% | 4,781 | 5,898 | 81.1% |
| Yorkshire & Humber | 3,574,600 | 4,271,381 | 83.7% | 4,081 | 4,856 | 84.0% |
| England | 34,897,648 | 43,644,520 | 80.0% | 40,162 | 49,461 | 81.2% |

Note: Population for England and deaths that occurred in England are for people 19 or over, whilst our cohort is limited to people aged between 19 and 100. Source: Population for England: Estimates of the population for the UK, England and Wales, Scotland and Northern Ireland (ONS); Deaths in England: Deaths registered weekly in England and Wales, provisional (ONS).

**Supplementary Table 2:** Performance of the risk models to predict risk of COVID-19 death in 14,104,452 patients registered with practices using the TPP System.

|  | Period 1<br>(24/01/2020 - 30/04/2020) |  | Period 2<br>(01/05/2020 - 28/07/2020) |  |
| --- | --- | --- | --- | --- |
|  | **COVID -19 death** | **COVID -19 death** | **COVID -19 death** | **COVID -19 death** |
|  | **females** | **males** | **females** | **males** |
|  | Estimate (95% CI) | Estimate (95% CI) | Estimate (95% CI) | Estimate (95% CI) |
| R<sup>2</sup> statistic | 0.778<br>(0.773 to 0.783) | 0.785<br>(0.780 to 0.790) | 0.769<br>(0.763 to 0.775) | 0.766<br>(0.760 to 0.772) |
| D statistic | 3.833<br>(3.775 to 3.892) | 3.911<br>(3.857 to 3.965) | 3.731<br>(3.669 to 3.793) | 3.704<br>(3.639 to 3.769) |
| Harrell's C statistic | 0.945<br>(0.942 to 0.948) | 0.934<br>(0.931 to 0.937) | 0.957<br>(0.953 to 0.960) | 0.948<br>(0.945 to 0.952) |
| Brier score | 0.0011 | 0.0015 | 0.0007 | 0.0008 |

**Supplementary Table 3:** Performance of the risk models to predict risk of COVID-19 death in the validation cohort by subgroup using Harrell’s C statistic (95% CI).

|  | Period 1 (24/01/2020 - 30/04/2020) |  | Period 2 (01/05/2020 - 28/07/2020) |  |
| --- | --- | --- | --- | --- |
|  | **COVID -19 death** | **COVID -19 death** | **COVID -19 death** | **COVID -19 death** |

|  | females | males | females | males |
| --- | --- | --- | --- | --- |
| **Age band** |  |  |  |  |
| 19-40 | 0.810 (0.753 to 0.866) | 0.833 (0.784 to 0.881) | 0.800 (0.676 to 0.925) | 0.786 (0.685 to 0.886) |
| 40-45 | 0.856 (0.798 to 0.913) | 0.811 (0.758 to 0.865) | 0.746 (0.659 to 0.833) | 0.763 (0.667 to 0.860) |
| 45-50 | 0.849 (0.808 to 0.890) | 0.841 (0.809 to 0.872) | 0.809 (0.723 to 0.895) | 0.768 (0.692 to 0.843) |
| 50-55 | 0.874 (0.847 to 0.901) | 0.834 (0.808 to 0.860) | 0.782 (0.716 to 0.849) | 0.779 (0.731 to 0.827) |
| 55-60 | 0.844 (0.819 to 0.869) | 0.796 (0.774 to 0.818) | 0.805 (0.754 to 0.856) | 0.809 (0.776 to 0.842) |
| 60-65 | 0.851 (0.831 to 0.872) | 0.808 (0.791 to 0.825) | 0.818 (0.777 to 0.858) | 0.788 (0.758 to 0.819) |
| 65-70 | 0.844 (0.826 to 0.861) | 0.803 (0.789 to 0.817) | 0.787 (0.755 to 0.819) | 0.817 (0.794 to 0.839) |
| 70-75 | 0.851 (0.837 to 0.865) | 0.796 (0.785 to 0.807) | 0.857 (0.838 to 0.876) | 0.826 (0.810 to 0.842) |
| 75-80 | 0.846 (0.835 to 0.857) | 0.808 (0.798 to 0.817) | 0.839 (0.824 to 0.855) | 0.821 (0.807 to 0.835) |
| 80-85 | 0.819 (0.810 to 0.828) | 0.787 (0.779 to 0.796) | 0.828 (0.816 to 0.840) | 0.803 (0.791 to 0.815) |
| 85-90 | 0.793 (0.784 to 0.802) | 0.768 (0.759 to 0.777) | 0.785 (0.774 to 0.797) | 0.755 (0.742 to 0.769) |
| 90+ | 0.736 (0.727 to 0.744) | 0.729 (0.719 to 0.740) | 0.750 (0.740 to 0.760) | 0.720 (0.705 to 0.735) |
| **Ethnicity** |  |  |  |  |
| Bangladeshi | 0.942 (0.918 to 0.966) | 0.939 (0.921 to 0.957) | 0.932 (0.891 to 0.973) | 0.918 (0.863 to 0.974) |
| Black African | 0.888 (0.859 to 0.917) | 0.906 (0.888 to 0.925) | 0.839 (0.743 to 0.935) | 0.908 (0.870 to 0.946) |
| Black Caribbean | 0.924 (0.912 to 0.937) | 0.930 (0.920 to 0.939) | 0.939 (0.914 to 0.965) | 0.921 (0.890 to 0.951) |
| Chinese | 0.960 (0.937 to 0.984) | 0.914 (0.877 to 0.950) | 0.927 (0.861 to 0.993) | 0.955 (0.913 to 0.997) |
| Indian | 0.939 (0.929 to 0.949) | 0.916 (0.906 to 0.926) | 0.929 (0.907 to 0.950) | 0.901 (0.878 to 0.924) |
| Mixed | 0.947 (0.923 to 0.972) | 0.972 (0.961 to 0.984) | 0.980 (0.966 to 0.994) | 0.963 (0.946 to 0.979) |
| Other | 0.935 (0.919 to 0.950) | 0.905 (0.892 to 0.918) | 0.883 (0.826 to 0.939) | 0.905 (0.880 to 0.930) |
| Pakistani | 0.918 (0.899 to 0.937) | 0.923 (0.908 to 0.937) | 0.948 (0.926 to 0.971) | 0.897 (0.865 to 0.930) |
| White British | 0.946 (0.944 to 0.948) | 0.935 (0.933 to 0.937) | 0.956 (0.954 to 0.958) | 0.947 (0.944 to 0.949) |
| White other | 0.963 (0.956 to 0.969) | 0.950 (0.943 to 0.957) | 0.968 (0.956 to 0.981) | 0.951 (0.939 to 0.963) |
| **Townsend quintile** |  |  |  |  |
| 1 (most affluent) | 0.945 (0.940 to 0.949) | 0.933 (0.929 to 0.937) | 0.961 (0.957 to 0.965) | 0.949 (0.944 to 0.954) |
| 2 | 0.945 (0.940 to 0.949) | 0.935 (0.932 to 0.939) | 0.955 (0.951 to 0.960) | 0.947 (0.942 to 0.952) |
| 3 | 0.947 (0.943 to 0.951) | 0.933 (0.929 to 0.937) | 0.958 (0.954 to 0.963) | 0.945 (0.940 to 0.950) |
| 4 | 0.946 (0.942 to 0.949) | 0.938 (0.935 to 0.942) | 0.956 (0.952 to 0.960) | 0.944 (0.939 to 0.950) |
| 5 (most deprived) | 0.943 (0.939 to 0.947) | 0.934 (0.930 to 0.937) | 0.952 (0.946 to 0.957) | 0.936 (0.930 to 0.942) |

**Supplementary Table 4:** D statistic of the risk models to predict risk of COVID-19 death the validation cohort by subgroup.

|  | Period 1 (24/01/2020 -30/04/2020) |  | Period 2 (01/05/2020 -28/07/2020) |  |
| --- | --- | --- | --- | --- |
|  | COVID -19 death | COVID -19 death | COVID -19 death | COVID -19 death |
|  | females | males | females | males |
| **Age band** |  |  |  |  |
| 19-40 | 2.175 (1.808 to 2.541) | 2.551 (2.192 to 2.910) | 2.376 (1.631 to 3.121) | 1.857 (1.230 to 2.484) |
| 40-45 | 2.537 (2.052 to 3.022) | 2.212 (1.826 to 2.598) | 1.360 (0.641 to 2.078) | 1.573 (0.942 to 2.204) |
| 45-50 | 2.660 (2.320 to 3.001) | 2.331 (2.053 to 2.610) | 1.966 (1.426 to 2.505) | 1.865 (1.401 to 2.329) |
| 50-55 | 2.627 (2.378 to 2.876) | 2.355 (2.144 to 2.566) | 1.961 (1.522 to 2.400) | 1.780 (1.462 to 2.099) |
| 55-60 | 2.434 (2.226 to 2.641) | 2.099 (1.943 to 2.255) | 1.946 (1.607 to 2.285) | 2.028 (1.773 to 2.283) |
| 60-65 | 2.603 (2.427 to 2.779) | 2.260 (2.129 to 2.390) | 2.003 (1.734 to 2.271) | 1.923 (1.713 to 2.133) |
| 65-70 | 2.433 (2.284 to 2.583) | 2.299 (2.185 to 2.414) | 1.904 (1.694 to 2.115) | 2.094 (1.918 to 2.269) |
| 70-75 | 2.735 (2.610 to 2.860) | 2.273 (2.182 to 2.365) | 2.668 (2.497 to 2.839) | 2.405 (2.268 to 2.541) |
| 75-80 | 2.609 (2.513 to 2.705) | 2.241 (2.168 to 2.314) | 2.454 (2.318 to 2.590) | 2.297 (2.187 to 2.408) |
| 80-85 | 2.304 (2.229 to 2.378) | 2.135 (2.071 to 2.199) | 2.285 (2.188 to 2.382) | 2.218 (2.123 to 2.312) |
| 85-90 | 2.011 (1.945 to 2.077) | 1.800 (1.739 to 1.861) | 1.945 (1.861 to 2.030) | 1.739 (1.652 to 1.826) |
| 90+ | 1.444 (1.385 to 1.503) | 1.446 (1.381 to 1.511) | 1.327 (1.263 to 1.392) | 1.347 (1.254 to 1.440) |
| **Ethnicity** |  |  |  |  |
| Bangladeshi | 3.370 (2.920 to 3.820) | 4.803 (4.345 to 5.261) | 3.666 (2.323 to 5.010) | 2.828 (2.073 to 3.582) |
| Black African | 3.560 (3.199 to 3.920) | 3.435 (3.196 to 3.675) | 2.648 (1.845 to 3.451) | 2.739 (2.247 to 3.232) |
| Black Caribbean | 3.973 (3.692 to 4.254) | 3.426 (3.259 to 3.594) | 3.670 (3.075 to 4.266) | 3.314 (2.922 to 3.705) |
| Chinese | 4.408 (3.753 to 5.063) | 4.730 (4.068 to 5.393) | 6.309 (4.459 to 8.160) | 4.571 (3.063 to 6.079) |
| Indian | 3.504 (3.304 to 3.704) | 3.338 (3.186 to 3.490) | 3.705 (3.289 to 4.121) | 2.826 (2.504 to 3.148) |
| Mixed | 4.039 (3.612 to 4.466) | 4.171 (3.818 to 4.525) | 5.267 (4.447 to 6.086) | 3.900 (3.336 to 4.464) |
| Other | 3.994 (3.701 to 4.286) | 3.725 (3.502 to 3.948) | 3.317 (2.786 to 3.848) | 3.070 (2.640 to 3.500) |
| Pakistani | 3.567 (3.240 to 3.895) | 3.381 (3.148 to 3.615) | 3.362 (2.832 to 3.892) | 2.623 (2.267 to 2.979) |
| White British | 3.487 (3.456 to 3.517) | 3.761 (3.729 to 3.792) | 3.595 (3.556 to 3.633) | 3.993 (3.945 to 4.041) |
| White other | 3.749 (3.614 to 3.884) | 4.007 (3.857 to 4.158) | 3.902 (3.667 to 4.137) | 4.223 (3.929 to 4.517) |

| **Townsend**<br>**quintile** |  |  |  |  |
| --- | --- | --- | --- | --- |
| 1 (most affluent) | 3.667 (3.591 to 3.744) | 3.738 (3.674 to 3.803) | 3.954 (3.863 to 4.045) | 4.253 (4.153 to 4.353) |
| 2 | 3.905 (3.826 to 3.985) | 3.875 (3.809 to 3.940) | 3.640 (3.558 to 3.722) | 3.686 (3.605 to 3.766) |
| 3 | 3.764 (3.690 to 3.838) | 4.006 (3.931 to 4.082) | 3.819 (3.727 to 3.911) | 3.562 (3.476 to 3.647) |
| 4 | 3.364 (3.304 to 3.424) | 4.070 (3.997 to 4.143) | 3.803 (3.706 to 3.899) | 3.664 (3.570 to 3.758) |
| 5 (most deprived) | 3.274 (3.218 to 3.330) | 3.920 (3.854 to 3.985) | 3.601 (3.509 to 3.693) | 3.696 (3.585 to 3.807) |

**Supplementary Table 5:** R-squared of the risk models to predict risk of COVID-19 death the validation cohort by subgroup.

|  | Period 1 (24/01/2020 -30/04/2020) |  | Period 2 (01/05/2020 -28/07/2020) |  |
| --- | --- | --- | --- | --- |
|  | COVID -19 death | COVID -19 death | COVID -19 death | COVID -19 death |
|  | females | males | females | males |
| **Age band** |  |  |  |  |
| 19-40 | 0.530 (0.438 to 0.607) | 0.608 (0.534 to 0.669) | 0.574 (0.388 to 0.699) | 0.451 (0.265 to 0.596) |
| 40-45 | 0.606 (0.501 to 0.686) | 0.539 (0.443 to 0.617) | 0.306 (0.089 to 0.508) | 0.371 (0.175 to 0.537) |
| 45-50 | 0.628 (0.562 to 0.683) | 0.565 (0.502 to 0.619) | 0.480 (0.327 to 0.600) | 0.454 (0.319 to 0.564) |
| 50-55 | 0.622 (0.574 to 0.664) | 0.570 (0.523 to 0.611) | 0.479 (0.356 to 0.579) | 0.431 (0.338 to 0.513) |
| 55-60 | 0.586 (0.542 to 0.625) | 0.513 (0.474 to 0.548) | 0.475 (0.382 to 0.555) | 0.495 (0.429 to 0.554) |
| 60-65 | 0.618 (0.584 to 0.648) | 0.549 (0.520 to 0.577) | 0.489 (0.418 to 0.552) | 0.469 (0.412 to 0.521) |
| 65-70 | 0.586 (0.555 to 0.614) | 0.558 (0.533 to 0.582) | 0.464 (0.407 to 0.516) | 0.511 (0.468 to 0.551) |
| 70-75 | 0.641 (0.619 to 0.661) | 0.552 (0.532 to 0.572) | 0.630 (0.598 to 0.658) | 0.580 (0.551 to 0.607) |
| 75-80 | 0.619 (0.601 to 0.636) | 0.545 (0.529 to 0.561) | 0.590 (0.562 to 0.616) | 0.558 (0.533 to 0.581) |
| 80-85 | 0.559 (0.543 to 0.574) | 0.521 (0.506 to 0.536) | 0.555 (0.533 to 0.575) | 0.540 (0.518 to 0.561) |
| 85-90 | 0.491 (0.475 to 0.507) | 0.436 (0.419 to 0.452) | 0.475 (0.453 to 0.496) | 0.419 (0.394 to 0.443) |
| 90+ | 0.332 (0.314 to 0.350) | 0.333 (0.313 to 0.353) | 0.296 (0.276 to 0.316) | 0.302 (0.273 to 0.331) |
| **Ethnicity** |  |  |  |  |
| Bangladeshi | 0.731 (0.671 to 0.777) | 0.846 (0.818 to 0.869) | 0.762 (0.563 to 0.857) | 0.656 (0.506 to 0.754) |
| Black African | 0.752 (0.710 to 0.786) | 0.738 (0.709 to 0.763) | 0.626 (0.448 to 0.740) | 0.642 (0.547 to 0.714) |
| Black Caribbean | 0.790 (0.765 to 0.812) | 0.737 (0.717 to 0.755) | 0.763 (0.693 to 0.813) | 0.724 (0.671 to 0.766) |
| Chinese | 0.823 (0.771 to 0.860) | 0.842 (0.798 to 0.874) | 0.905 (0.826 to 0.941) | 0.833 (0.691 to 0.898) |
| Indian | 0.746 (0.723 to 0.766) | 0.727 (0.708 to 0.744) | 0.766 (0.721 to 0.802) | 0.656 (0.600 to 0.703) |
| Mixed | 0.796 (0.757 to 0.826) | 0.806 (0.777 to 0.830) | 0.869 (0.825 to 0.898) | 0.784 (0.727 to 0.826) |
| Other | 0.792 (0.766 to 0.814) | 0.768 (0.745 to 0.788) | 0.724 (0.649 to 0.780) | 0.692 (0.625 to 0.745) |
| Pakistani | 0.752 (0.715 to 0.784) | 0.732 (0.703 to 0.757) | 0.730 (0.657 to 0.783) | 0.622 (0.551 to 0.679) |
| White British | 0.744 (0.740 to 0.747) | 0.771 (0.768 to 0.774) | 0.755 (0.751 to 0.759) | 0.792 (0.788 to 0.796) |
| White other | 0.770 (0.757 to 0.783) | 0.793 (0.780 to 0.805) | 0.784 (0.762 to 0.803) | 0.810 (0.787 to 0.830) |

| **Townsend**<br>**quintile** |  |  |  |  |
| --- | --- | --- | --- | --- |
| 1 (most affluent) | 0.763 (0.755 to 0.770) | 0.769 (0.763 to 0.775) | 0.789 (0.781 to 0.796) | 0.812 (0.805 to 0.819) |
| 2 | 0.785 (0.777 to 0.791) | 0.782 (0.776 to 0.787) | 0.760 (0.751 to 0.768) | 0.764 (0.756 to 0.772) |
| 3 | 0.772 (0.765 to 0.779) | 0.793 (0.787 to 0.799) | 0.777 (0.768 to 0.785) | 0.752 (0.743 to 0.760) |
| 4 | 0.730 (0.723 to 0.737) | 0.798 (0.792 to 0.804) | 0.775 (0.766 to 0.784) | 0.762 (0.753 to 0.771) |
| 5 (most deprived) | 0.719 (0.712 to 0.726) | 0.786 (0.780 to 0.791) | 0.756 (0.746 to 0.765) | 0.765 (0.754 to 0.776) |

**Supplementary Table 6.**
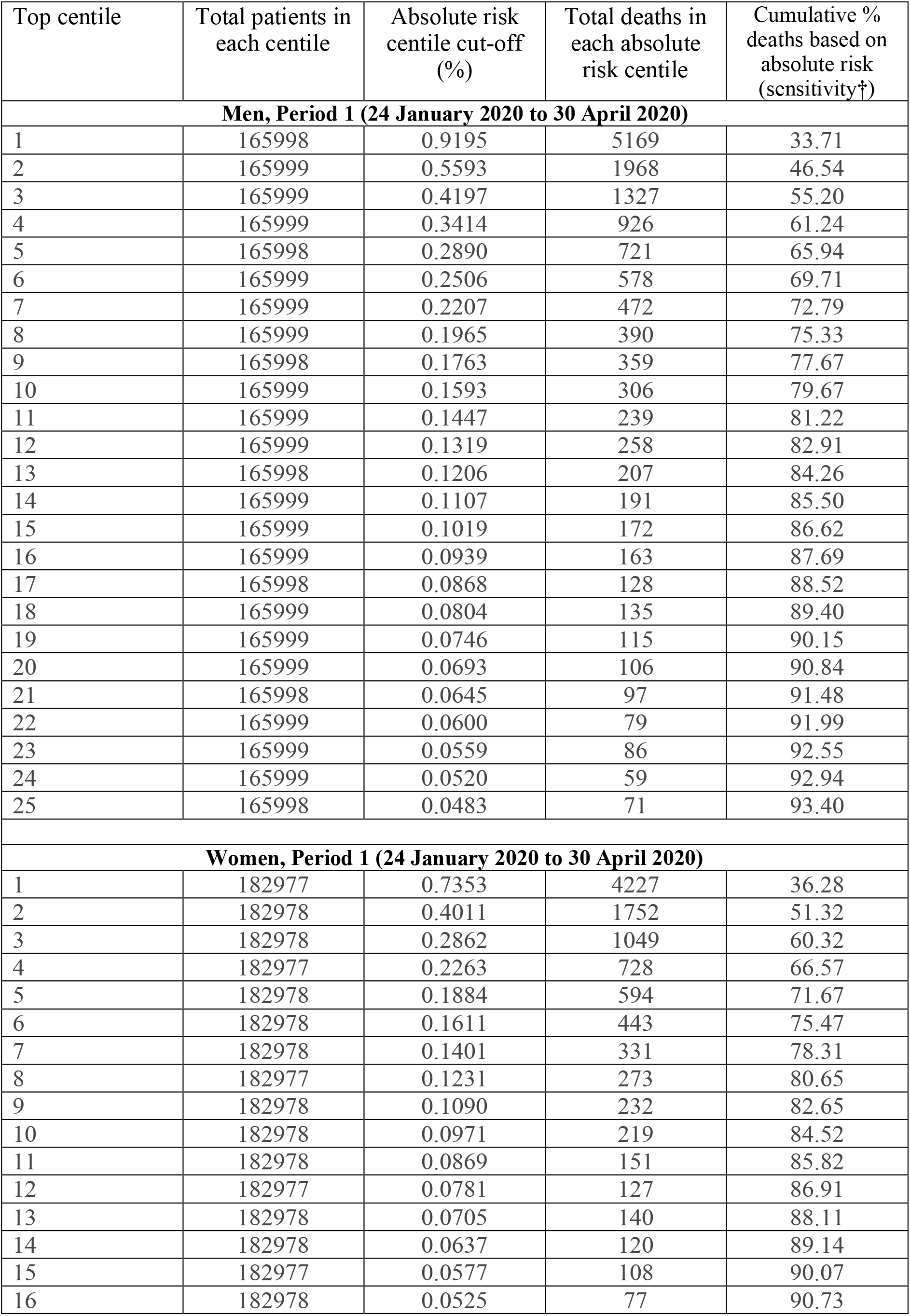
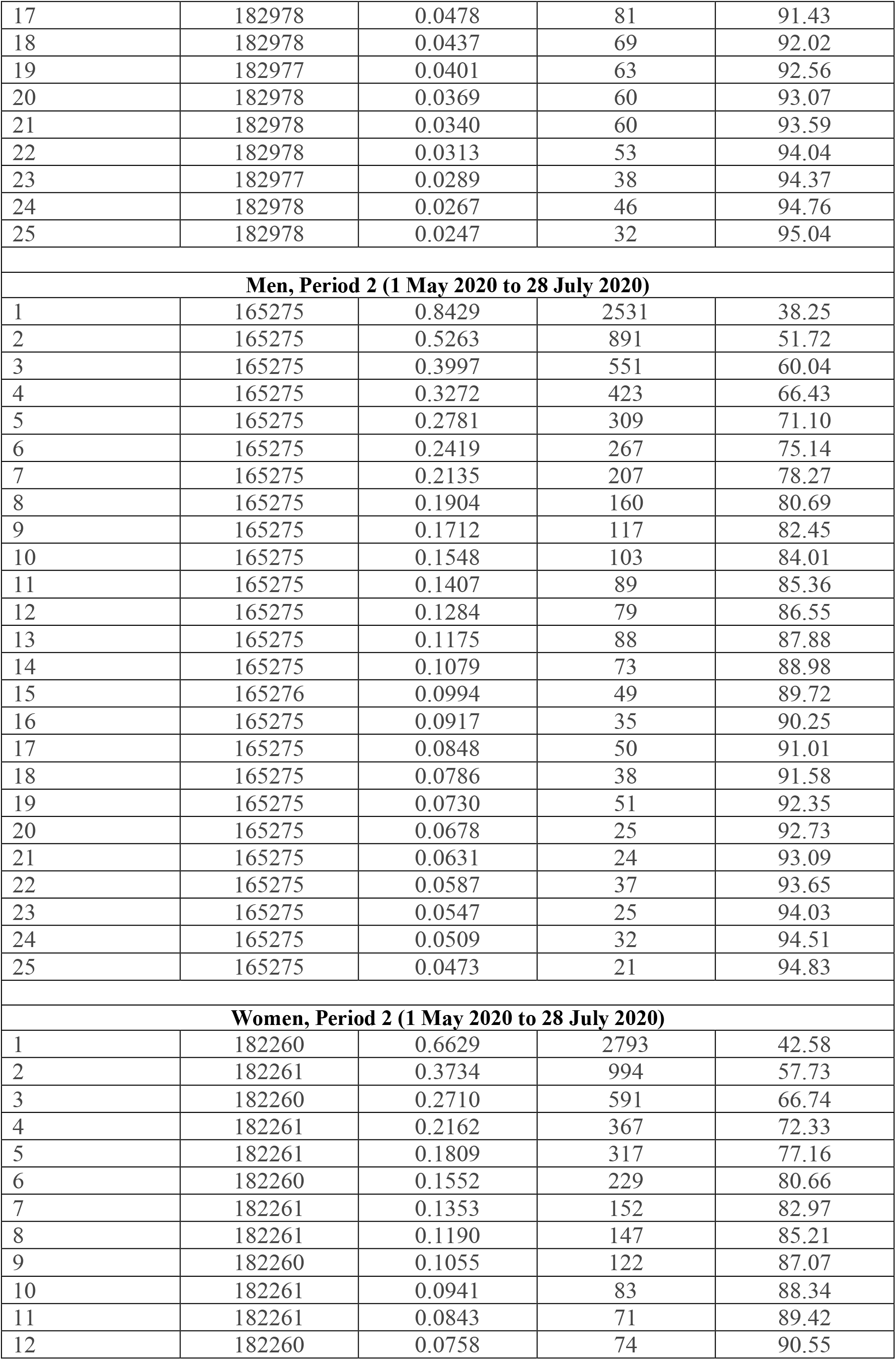
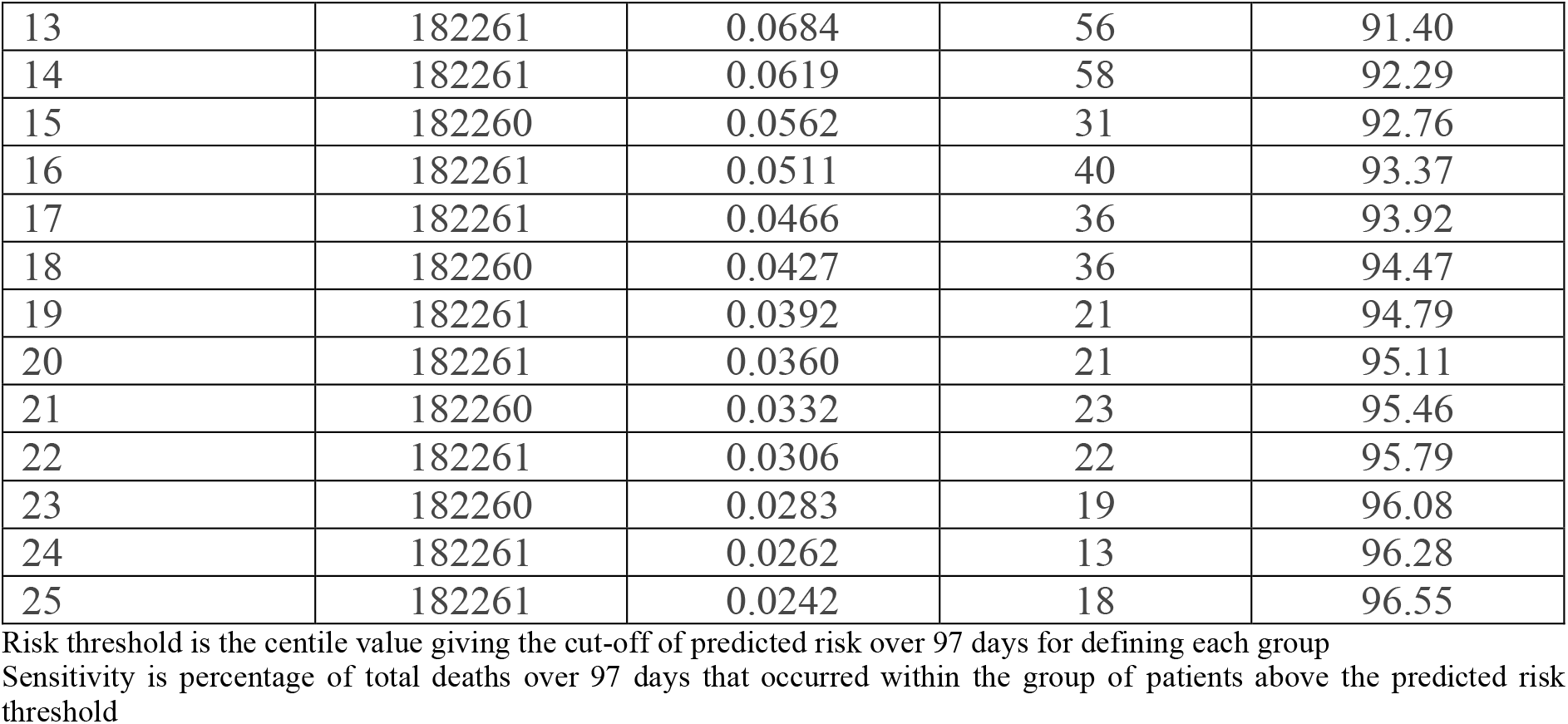
Sensitivity for covid-19 related death by sex at different absolute risk thresholds.

| Top centile | Total patients in each centile | Absolute risk centile cut-off (%) | Total deaths in each absolute risk centile | Cumulative % deaths based on absolute risk (sensitivity†) |
| --- | --- | --- | --- | --- |
| **Men, Period 1 (24 January 2020 to 30 April 2020)** |  |  |  |  |
| 1 | 165998 | 0.9195 | 5169 | 33.71 |
| 2 | 165999 | 0.5593 | 1968 | 46.54 |
| 3 | 165999 | 0.4197 | 1327 | 55.20 |
| 4 | 165999 | 0.3414 | 926 | 61.24 |
| 5 | 165998 | 0.2890 | 721 | 65.94 |
| 6 | 165999 | 0.2506 | 578 | 69.71 |
| 7 | 165999 | 0.2207 | 472 | 72.79 |
| 8 | 165999 | 0.1965 | 390 | 75.33 |
| 9 | 165998 | 0.1763 | 359 | 77.67 |
| 10 | 165999 | 0.1593 | 306 | 79.67 |
| 11 | 165999 | 0.1447 | 239 | 81.22 |
| 12 | 165999 | 0.1319 | 258 | 82.91 |
| 13 | 165998 | 0.1206 | 207 | 84.26 |
| 14 | 165999 | 0.1107 | 191 | 85.50 |
| 15 | 165999 | 0.1019 | 172 | 86.62 |
| 16 | 165999 | 0.0939 | 163 | 87.69 |
| 17 | 165998 | 0.0868 | 128 | 88.52 |
| 18 | 165999 | 0.0804 | 135 | 89.40 |
| 19 | 165999 | 0.0746 | 115 | 90.15 |
| 20 | 165999 | 0.0693 | 106 | 90.84 |
| 21 | 165998 | 0.0645 | 97 | 91.48 |
| 22 | 165999 | 0.0600 | 79 | 91.99 |
| 23 | 165999 | 0.0559 | 86 | 92.55 |
| 24 | 165999 | 0.0520 | 59 | 92.94 |
| 25 | 165998 | 0.0483 | 71 | 93.40 |
| **Women, Period 1 (24 January 2020 to 30 April 2020)** |  |  |  |  |
| 1 | 182977 | 0.7353 | 4227 | 36.28 |
| 2 | 182978 | 0.4011 | 1752 | 51.32 |
| 3 | 182978 | 0.2862 | 1049 | 60.32 |
| 4 | 182977 | 0.2263 | 728 | 66.57 |
| 5 | 182978 | 0.1884 | 594 | 71.67 |
| 6 | 182978 | 0.1611 | 443 | 75.47 |
| 7 | 182978 | 0.1401 | 331 | 78.31 |
| 8 | 182977 | 0.1231 | 273 | 80.65 |
| 9 | 182978 | 0.1090 | 232 | 82.65 |
| 10 | 182978 | 0.0971 | 219 | 84.52 |
| 11 | 182978 | 0.0869 | 151 | 85.82 |
| 12 | 182977 | 0.0781 | 127 | 86.91 |
| 13 | 182978 | 0.0705 | 140 | 88.11 |
| 14 | 182978 | 0.0637 | 120 | 89.14 |
| 15 | 182977 | 0.0577 | 108 | 90.07 |
| 16 | 182978 | 0.0525 | 77 | 90.73 |
| 17 | 182978 | 0.0478 | 81 | 91.43 |
| 18 | 182978 | 0.0437 | 69 | 92.02 |
| 19 | 182977 | 0.0401 | 63 | 92.56 |
| 20 | 182978 | 0.0369 | 60 | 93.07 |
| 21 | 182978 | 0.0340 | 60 | 93.59 |
| 22 | 182978 | 0.0313 | 53 | 94.04 |
| 23 | 182977 | 0.0289 | 38 | 94.37 |
| 24 | 182978 | 0.0267 | 46 | 94.76 |
| 25 | 182978 | 0.0247 | 32 | 95.04 |
| **Men, Period 2 (1 May 2020 to 28 July 2020)** |  |  |  |  |
| 1 | 165275 | 0.8429 | 2531 | 38.25 |
| 2 | 165275 | 0.5263 | 891 | 51.72 |
| 3 | 165275 | 0.3997 | 551 | 60.04 |
| 4 | 165275 | 0.3272 | 423 | 66.43 |
| 5 | 165275 | 0.2781 | 309 | 71.10 |
| 6 | 165275 | 0.2419 | 267 | 75.14 |
| 7 | 165275 | 0.2135 | 207 | 78.27 |
| 8 | 165275 | 0.1904 | 160 | 80.69 |
| 9 | 165275 | 0.1712 | 117 | 82.45 |
| 10 | 165275 | 0.1548 | 103 | 84.01 |
| 11 | 165275 | 0.1407 | 89 | 85.36 |
| 12 | 165275 | 0.1284 | 79 | 86.55 |
| 13 | 165275 | 0.1175 | 88 | 87.88 |
| 14 | 165275 | 0.1079 | 73 | 88.98 |
| 15 | 165276 | 0.0994 | 49 | 89.72 |
| 16 | 165275 | 0.0917 | 35 | 90.25 |
| 17 | 165275 | 0.0848 | 50 | 91.01 |
| 18 | 165275 | 0.0786 | 38 | 91.58 |
| 19 | 165275 | 0.0730 | 51 | 92.35 |
| 20 | 165275 | 0.0678 | 25 | 92.73 |
| 21 | 165275 | 0.0631 | 24 | 93.09 |
| 22 | 165275 | 0.0587 | 37 | 93.65 |
| 23 | 165275 | 0.0547 | 25 | 94.03 |
| 24 | 165275 | 0.0509 | 32 | 94.51 |
| 25 | 165275 | 0.0473 | 21 | 94.83 |
| **Women, Period 2 (1 May 2020 to 28 July 2020)** |  |  |  |  |
| 1 | 182260 | 0.6629 | 2793 | 42.58 |
| 2 | 182261 | 0.3734 | 994 | 57.73 |
| 3 | 182260 | 0.2710 | 591 | 66.74 |
| 4 | 182261 | 0.2162 | 367 | 72.33 |
| 5 | 182261 | 0.1809 | 317 | 77.16 |
| 6 | 182260 | 0.1552 | 229 | 80.66 |
| 7 | 182261 | 0.1353 | 152 | 82.97 |
| 8 | 182261 | 0.1190 | 147 | 85.21 |
| 9 | 182260 | 0.1055 | 122 | 87.07 |
| 10 | 182261 | 0.0941 | 83 | 88.34 |
| 11 | 182261 | 0.0843 | 71 | 89.42 |
| 12 | 182260 | 0.0758 | 74 | 90.55 |
| 13 | 182261 | 0.0684 | 56 | 91.40 |
| 14 | 182261 | 0.0619 | 58 | 92.29 |
| 15 | 182260 | 0.0562 | 31 | 92.76 |
| 16 | 182261 | 0.0511 | 40 | 93.37 |
| 17 | 182261 | 0.0466 | 36 | 93.92 |
| 18 | 182260 | 0.0427 | 36 | 94.47 |
| 19 | 182261 | 0.0392 | 21 | 94.79 |
| 20 | 182261 | 0.0360 | 21 | 95.11 |
| 21 | 182260 | 0.0332 | 23 | 95.46 |
| 22 | 182261 | 0.0306 | 22 | 95.79 |
| 23 | 182260 | 0.0283 | 19 | 96.08 |
| 24 | 182261 | 0.0262 | 13 | 96.28 |
| 25 | 182261 | 0.0242 | 18 | 96.55 |

Risk threshold is the centile value giving the cut-off of predicted risk over 97 days for defining each group
Sensitivity is percentage of total deaths over 97 days that occurred within the group of patients above the predicted risk threshold

**Supplementary Figure 1.**
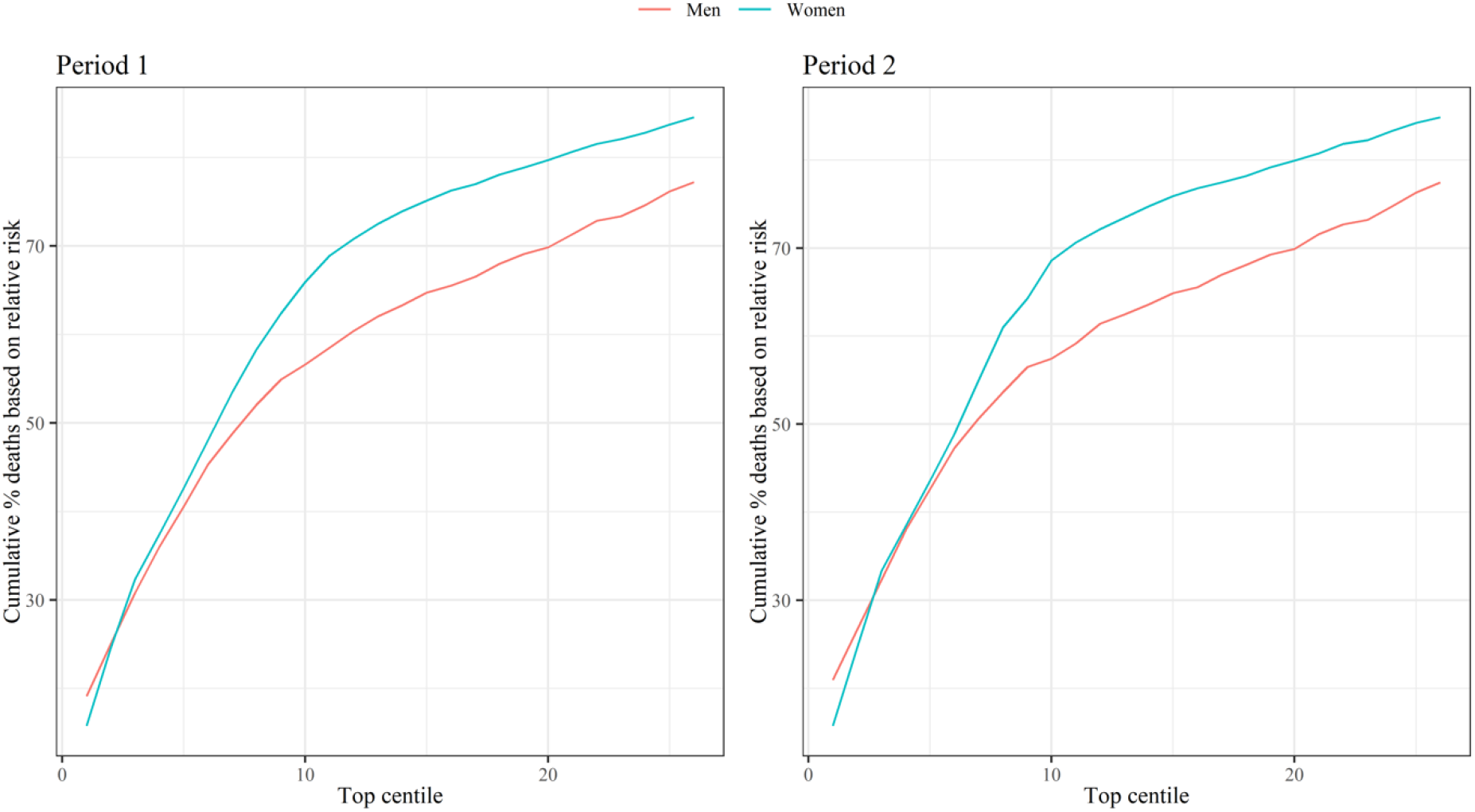
Centiles based on predicted relative risks compared with someone of the same age/sex with no risk factors. Sensitivity is percentage of total deaths in the period that occurred within the group of patients above the predicted risk threshold.

